# Evaluating the implementation outcome “acceptability” of the Eight (8) Antenatal Care contacts, and modelling the predictors that determine compliance by women attending Antenatal care clinics in Wakiso district

**DOI:** 10.1101/2023.01.06.23284275

**Authors:** Innocent Ssemanda, Kalembe Brenda, Riri Johnson Vonje, Karen Mwengwe, Okwadi Tukei, Oluseye A Ogunbayo

## Abstract

**Background:** The World Health Organization increased the number of antenatal care (ANC) visits from Four (4) to Eight (8) visits or more, to reduce the exponentiated incidences of preventable deaths of newborns, maternal mortality, perinatal, and stillbirths. Unfortunately, previous studies have indicated that pregnant women are noncompliant and nonadherence to the number of antenatal visits recommended by WHO. Therefore, the study measured the level of acceptability of the Eight (8) antenatal care contacts and determined the predictors’ variable that influenced compliance to 8-ANC visit in women attending ANC in the Wakiso district.

**Methodology:** This was a hospital-based medical records observational-cross-section study design, that used a multi-stage-stratified sampling technique to collect data from eligible women, Antenatal Progressive examination cards, and ANC registers, after obtaining ethical approval. A questionnaire survey and checklist were used to collect the data on the acceptability of the 8-ANC from January to April 2022. 401 eligible full-term pregnant and postnatal women were invited to participate in the study, and Informed consent was obtained The data collected was cleaned, coded in Microsoft Excel software, and imported into STATA version 15 for analysis. Q-Q-plot and histogram were used to check the normality assumption of the data. Descriptive statistics were reported using a t-test. Aiken Information Criterion (AIC) and Bayesian Information Criterion (BIC) measures were used to select the best fit model, then the inferential statistics were reported after using a generalized leaner for the poison model. The level of significance was set at P < 0.05.

**Results:** 401 eligible full-term pregnant and postnatal women participated in the study, out of which 101(25.2%) were postnatal women 300(74.8%) were full-term pregnant women and they had a mean age of 24.8 years, with an SD of 6.31 years. 193(48.13%) were married while 208(51.87) were not married. The average number of ANC visits a pregnant could accept to complete was 4 contacts. The level of acceptability to complete the 8-ANC visits was 27(6.73%) among the primigravida group at estimate of (-.222, at 95% CI [-.328 -.116], P=0.001), and 19(4.74%) among the multigravida group at estimate of 2.04 %, 95% CI [3.811, 4.184], *P*-value = 0.001. The predictor variables of the acceptability of 8-ANC visits were health system delay at ANC was .0384%, 95% CI; (-.073, -0343), *P*-value= 0.003), gestation age at which ANC started.153%, 95%; (-.252 -.054), *P-* value *=*0.002). The level of income a woman has2.025%, 95% CI; (3.001 1.047), *P-*value *=*0.001), and the level of awareness about the completion of the the8-ANC visit 1.413, 95%, CI; (1.998 3.828), *P-*value*=*0.001).

**Conclusion:** The level of acceptability to complete the 8-ANC visits or more was Low at Kasangati health centre IV, and this was influenced by; health system delay at ANC, high gestation age’(delay to start ANC service), level of income, and lack of awareness about the completion of the 8-ANC visits. These factors should be addressed in the social community to scale-up acceptability to complete the 8-ANC contacts or more. Among full-term pregnant and postnatal women in Wakiso District

## Introduction

## Background

Antenatal care is the type of healthcare service given to pregnant women till labour by health professionals. In 2016, the World Health Organization introduced the Eight(8) antenatal care visits model(8-ANC) to replace the traditional Four(4) times antenatal care contacts(4-ANC)(1). This was done to achieve the United Nations sustainable development goal 3(three) of good health and well-being targeting ending preventable newborn deaths by 2030(2, 3). In this, WHO recommends that contacts 8 or more pregnant women should attend ANC for the whole pregnancy course(1) Despite these tremendous global efforts, pregnant women in Low and middle-income countries (LMICs) have failed to accept to complete all 8-ANC visits(4). This failure is mostly observed in Uganda’s social communities(4, 5), no wonder why maternal and newborn mortality rates are still skyrocketing in some districts of the country(6). As indicated from previous literature, rates of maternal and newborn mortality correlate with the incompletion and noncompliance of these groups to the 8-ANC visit model(4, 7, 8), which signify that unacceptability, and underutilization of the Eight(8) antenatal care visits in LMICs is at prime(4, 9).

For example McNellan et al, 2019 Indicated that noncompliance and inadherence were associated with the acceptability of the Eight (8) Antenatal care visits model(10). Furthermore, acceptability and utilization of the 8-antenatal care visits were reported to have a strong correlation to the completion of the 8-ANC visits(11).

Globally, the impact of acceptability, utilization, and adoption of the Eight (8) antenatal care visits is negatively felt. For example, literature shows that globally it is estimated that over 2.4 million children die in their first 28 days of life in 2020(12), this signifies that approximately 6,500(0.03%) neonatal deaths get registered every day(13) in Africa, and Sub-Saharan Africa registered the highest neonatal mortality rate in 2019 at a rate of 27 deaths per 1,000 live births(14). In 2019, it is estimated that the neonatal mortality rate was 20 deaths per 1,000 live births in Uganda, and in Wakiso district where the study was conducted, it is estimated that more than 15 deaths per 1000 live birth occur annually(15). These death rates were significantly correlated with nonadherence, incompletion, and noncompliance to the 8-ANC visits or more among women attending ANC(4, 16). A current study done in the Uganda-Lira district indicated that maternal motility rate, perinatal death rate, and other obstetrics complications are unacceptably higher than expected among the pregnant women who are underutilized and are noncompliant with the 8-antenatal care visits recommended by WHO(17). Furthermore, Odusina et al, 2021 established a significant relationship between the completion of the Eight (8) antenatal care visits, and maternal, and newborn death rates(4), unfortunately, less was done to establish the level of acceptability of the 8-ANC visits. Additionally, previous literature indicated that to accelerate progress towards the reduction of newborn deaths, all pregnant women should be motivated to accept to attend ANC up to 8 contacts or more ANC visits during pregnancy(18). Furthermore, Okedo et al, 2019, conducted a systematic review and established the determinants of antenatal care utilization(19) which should be recognized as tremendous contributions to the completion of the 8-ANC visits. However, the same was supposed to be replicated by using different methodological approaches using primary data collected from different population in inductive-descriptive research methods. Literature further, have described the factors correlated with inadherence and noncompliance of pregnant women to the Eight (8) antenatal care visits(4, 6, 18, 20, 21). Unfortunately, little is done to assess the level of acceptability of the Eight (8) antenatal care visits or more among pregnant women in Uganda and literature that exist they enclose population, evidence and methodological research gaps, furthermore the studies that have evaluated the factors that influence the acceptability of the 8-ANC contacts are scanty in Uganda. Therefore, in this current study, we hypothesized that the level of acceptability of the 8-ANC visit or more is low, and its not associated with the completion of all the 8-ANC visits. Therefore, the study measured the level of acceptability of the 8-ANC visits, and its predictors to its acceptablity of the 8-ANC contact or more among pregnant women in the Wakiso district.

## Methods

### Study design and population

We conducted a hospital-based medical record observational-cross-section study design to collect data from eligible women who attended Kasanagti Health Centre IV-Antenatal care clinic (KHC-IV) and at Cleveland women and children’s specialized Clinic (CWCSC) in Wakiso district. The postnatal and full-term pregnant women were the eligible study participants, and they were recruited in the study from 15th January to 30^th^ April 2022.

### Study setting

The study was conducted in an uncontrolled environment at Kasanagti Health Centre IV (KHC-IV) and at Cleveland women and children’s specialized Clinic (CWCSC) in Wakiso district, where eligible women were invited to participate in the study which started from January to April 2022. Participants registered in the Antenatal care Register were phone called to come with their antenatal progressive examination cards if they are agreeable to participate in the study. A hospital Board room with a capacity to accommodate 200 people was used to sit the participants, and refreshing drinks were served, and

### The study area

Wakiso district has an estimated total population of about 2.5 million people out of which 1.5 are females(15). In the Wakiso district, 104 healthcare facilities are providing Antenatal care services, and it is estimated that Wakiso has the highest fertility rate of 50%, and it is reported that Wakiso District has the highest neonatal, infant, and maternal mortality rates in the country although it’s an urban(22). Wakiso district is a malaria-endemic area, and with increased HIV/AIDS infection prevalence, the majority of the population generally lives in absolute poverty, their main economic activities are agriculture and cattle rearing. The study assumed studying such a community with such characteristics and exposures shall generate data related to the outcome of our interest.

### Data collection tools and process

The data was collected using a questionnaire survey and a checklist we designed. The questionnaire survey was wrapped in an envelope and distributed to the participants who attended the study. Antenatal progressive examination cards were collected from the participants and distributed with a checklist to the researchers. A chart review was systematically done by the trained senior midwives and Gynecologists together with the assessed participant The data collected from the antenatal progressive examination card was matched with that recorded in the Antenatal care register, to assure its validity and reliability. Completeness and incompleteness of the Eight (8) or more ANC contacts were confirmed if the data on the examination card matches with that recorded in ANC Register. Furthermore, senior gynecologists gently palpated the abdomen with both the left and the right hand to determine the height of the uterine fundus of the uterus to match with the pregnancy gestation age versus the number of antenatal care visits completed, and this estimate was guided by Impey and friends(23). Completion of Eight (8) or more antenatal care contacts were defined as when a pregnant woman attended at least Eight(8) or more contacts to the ANC clinic for antenatal care services, and the gestation age was defined as the number of weeks the intrauterine fetus had at that particular time.

### Participants

The study participants were the women attending Antenatal Care Clinic (ANC) services at Kasangati Health Centre IV, (KHC IV) and at Cleveland women and children’s Specialized Clinic in Wakiso district.

### Inclusion criteria

The study considered only full-term pregnant women who are in their Third trimester, and those on postnatal care, at Kasangati Health Centre IV ANC clinic.

### Exclusion criteria

All pregnant women who were in their First and Second trimesters and those who had passed their postnatal period of Six weeks were excluded from the study.

### Sampling technique and procedures of participant selection

We used a multi-stage approach with a stratified sampling technique. In the initial stage, the stratification was done on the four sub-counties in the Wakiso district. The Two sub-counties were selected from the Four wards using a simple random sampling technique. In the second stage, Health centres found in the selected sub-counties were stratified into two categories that are “urban or rural area”. By purposively, the urban region was selected based on the fact that they have active ANC clinics, with high tune-ups of pregnant women attending ANC clinic services. Out of Eight (8) healthcare facilities with ANC clinics, two were selected by systematic random sampling technique. Then, a complete enumeration was done of every eligible participant found in the strata

### The study variable selection and measurement

The measure of acceptability of Eight (8) or more ANC contacts was determined by counting the absolute number of ANC contacts that appeared on both, the progressive examination card and in the ANC register, pregnant women attended at ANC clinic until birth. Furthermore, acceptability was measured based on the participants’ opinions and perceptions towards the completion of the 8-ANC contacts. For example, participants were asked to give their opinion on the acceptability to complete the 8-ANC contacts or more by stating either; strongly agree, agree, strongly disagree, or disagree to have the 8-ANC contacts or more. The exogenous variables like the attitude toward health work, Midwifes’ motivation, awareness about 8-ANC, Level of education, Level of income at home, being a Multigravida, being a primigravida, Gestation age in weeks, waiting time, Age of a woman and distance to the nearest ANC clinic were examined by asking the respondent to specify their level of agreement and satisfaction on 5-point rating scales. The responses generated were compared against acceptability to complete the 8-ANC contacts. The predictor variables: The study variable was based on the characteristics of the study subjects and the literature reviews; for example (4, 7, 9, 18, 19, 21, 24, 25),(26-28), and all these variables are assessed and recorded in the Integrated antenatal register and Antenatal progressive examination cards. The variables considered and included in the analysis were; the age of women, distance to the ANC clinic, parity, gravidity, waiting time in the queue, and Gestation age the ANC started were treated and collected as continuous data. While the variable “attitude of health workers, awareness about 8-ANC visits, level of income, and midwife-motivation were measured on the 3-point rating scale. For example; variable attitude was assessed as; 1 poor, 2 fair, and 3 good. The importance of 8-ANC visits among primigravida and multigravida was assessed as; 1 not important, 2 moderately important, and 3 very important. Further, pregnant women were asked how the level of income, and midwife’s motivation is likely to influence the completion of 8-ANC visits, and the responses were coded as; 1 not likely, 2 somewhat likely, and 3 very likely.

### Outcome variable

The outcome variable was the level of acceptability of the 8-–ANC contacts or more. This was collected as counts “the number of ANC contacts a pregnant woman has attended or completed. And these counts ranged between ANC contact 1 to 8-ANC contacts or more(29).

### Exposures

In the exposed group were the women in their third trimester; but at gestation age between 36-41 weeks of amenorrhea, who attended the monthly ANC clinic services, and they were assessed by counting the number of ANC visits attended. The unexposed group; was the women in the first, second, and third trimesters at gestation age ranging between 13 weeks to 30 weeks of Amenorrhea, and those who have passed their postnatal period were excluded from the study. In the study, the assumed potential confounding variable was controlled primarily during data collection; by randomization, and also during analysis, by fitting a regression model.

## Data

### Sources /measurement

Data were collected from Integrated Antenatal Register (HMIS FORM 071), Antenatal Progressive examination Cards which had information on ANC contacts attendance, and from full-term pregnant women and postnatal women. The study examined the ANC records starting from Dec 2021 to Aug 2022, and all the eligible females aged between 14-45 years who were full-term pregnant and those who were postnatal were considered to participate in the study. To get the number of ANC contacts, the age of the women, parity, gravidity, and gestation age that ANC contacts started; were collected from Integrated Antenatal Register (HMIS071), and the Antenatal progressive examination card. For the variable age was recorded as an absolute value. ANC contacts were measured as the number of times a pregnant woman visited the clinic for ANC services. Parity was the number of live children the woman has, and gravidity was assessed as the number of times that a woman has been pregnant. The gestation age at which ANC started was assessed based on the duration of pregnancy dated from the first day of the last menstrual period, and it was measured in weeks. The waiting time in the queue was assessed by asking the participants to report the amount of time waited in the queue before being attended to by a healthcare professional/ midwife. The variable distance in kilometers to the ANC clinic was also reported by the participants surveyed. To assess the Attitude of health workers, an attitude scale rating was used., to assess midwife motivation, level of income, and Awareness about 8-ANC visits. A 3-point rating scale was employed. For example, participants were asked to give their score concerning each statement on the 3-point rating scale.

### Sample size determination

The sample size was determined using a single sample size for the mean formula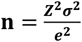. Where “**n**” is the sample size required, ***Z***^**2**^is the desired level of confidence 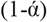 set at 1.96, at 95% confidence interval (CI), and ***e***^**2**^is the acceptable sampling error, set at ±5, **σ**^**2**^is the standard deviation set at 51.1. ***e***^**2**^&**σ**^**2**^Values were obtained from the pilot sample of 100 participants conducted at the health facility. Therefore, a minimum sample size of **401** participants was obtained and gave 80% power to detect an effect size(30). Therefore, the standard 10% of non-response rate was ignored.

### Study variables and how they were handled in the analysis

The outcome variable was the level of acceptability in terms of the number of ANC contacts s a pregnant woman has completed in the whole pregnancy course till birth, and it was handled “as a count”.Level of education, primigravida, multigravida, awareness about 8-ANC contacts, level of income, midwife motivation, and attitude of health workers were handled as categorical variables. The variables Age, distance to the ANC clinic, waiting time in the queue to be served by a midwife, parity gravidity, and gestation age at which ANC started were handled as continuous variables. The data collected from 401 self-administered questionnaire surveys, and chart review was checked for completeness, deleting the missing values, and unlikely responses were manually cleaned up on such indications. Data were extracted and entered directly into a computer Microsoft excel sheet, for systematic double-checking, cleaning, and coding. The data was imported into STATA version 15.1 (Stata-Corp, College Station, TX), where it was cross-checked for consistency, and accuracy, and analyzed.

### Data processing and analysis

For the descriptive statistics, the mean and standard deviation (SD) were reported for continuous variables, and frequency and percentages were reported for categorical variables. The inference statistics were reported using regression coefficient and confidence intervals (CI). A generalized leaner model for poison model (GLM) was fitted to determine the predictor variables that influenced acceptability to the completion of 8-ANC visits among pregnant and/or postnatal women. The best fit model was selected using Adjusted R-squared (R^2^), and by using the AIC and BIC measure. The Adjusted R^2^ from model 1 had 0.096 variations, while model 2 from stepwise regression had 0.102 variations in the outcome is accounted for by the explanatory variables. Comparing the two statistical models, either the Aiken Information Criterion (AIC) or Bayesian Information Criterion (BIC) criteria, the values from the stepwise regression were smaller; had 1614 values compared to the 1621 values from model 1.

This suggested that model 1 has the highest proportion of 102% of the variation being accounted for by the stepwise regression model, which had the lowest value of AIC and BIC, making it to be the best fit model. Further, the likelihood ratio test was done to select the model without interaction effect, at p-value=0.630. The Residual for linear regression was approximately normal with mean zero and constant variance. To have a prediction and precision of the prediction multicollinearity was checked using variance inflation factor (VIF) after fitting the regression model and all the variables had VIF between 1 to 4 suggesting that there is moderate correlation and it does not warrant corrective measures. Before fitting the model, checking for missing data in the dataset was done and indicated missing value =0(zero). Then a stepwise regression model was fit to compare the predictors against the dependent variable. The variables with p-value <0.05 in the model were considered independent predictor variables for the acceptability to completion of the 8-ANC visits, with a 95% confidence interval (CI) reported. During analysis, a multilevel generalized linear regression model (GLM) was fit to see through confounding factors and isolate the relationship of interest.

### Ethics statement

This study was approved by the medical board of Kasanagti Health centre IV, Uganda National council for science and technology (UNCST) Reg No. HS6997ES, and Indian Institute of Health and Allied Science Uganda (Reg No. IIASREC/0181). Hospital directors and district health officers were informed about the study ahead of time and informed consent was obtained. All participants were provided with written consent earlier to consent for their participation in this study and were aware and informed of their right to withdraw at any time. Participants travelled a long distance of about 2 kilometres and above, their transport fees were refunded, and they were also provided with light refreshments during the interview and discussion.

## Results

### Demographic characteristics

Table 1 is the demographic characteristics of the study participants and indicates that there were 401 women both full-term pregnant women and postnatal women participated in the study, of which 101(25.2%) were postnatal women, and 300(74.8%) were full-term pregnant women, and they had a mean age of 24.8 years, with the SD of 6.31 years. The study participants had a mean number of 2(Two) parity (P_2_) with the SD of 2(Two), and a mean number of gravidity 3(three) with the SD of 2(Two) pregnancies. In this region, pregnant women move a mean distance of 3.9 Kilometres to reach the nearest ANC clinic, with a SD distance of 1.8 Km. The proportion of women who were married and stayed with their husbands was 193(48.13%), while 208(51.87) were cohabiting with their partners. Out of the 401, women who participated in the study; 173(43.14%) were under certificate level, 20(29.93%) were at certificate level, and 108(26.93%) were at degree level. Among those participated in the study 122(30.424%) were Muslims, 106(26.433%) were Christian and 173(43.14%) were not sure of their religion

**Table 1.** Background characteristics of the study participants at Kasanagati Health centre IV, in Wakiso District.

| The summary statistics of the postnatal and pregnant women who participated in the study (obs =401) |  |  |  |  |
| --- | --- | --- | --- | --- |
| Characteristics | Min | Max | Mean (SD) | Proportion (%) |
| Age | 13 | 41 | 24.8 (6.31) |  |
| Distance to ANC clinic | 1 | 9 | 3.9(1.8) |  |
| Parity | 0 | 8 | 2(1.9) |  |
| Gravidity | 0 | 8 | 3(2.1) |  |
| Marital status (n=401) |  |  |  |  |
| Yes ( stay with husband) |  |  |  | 193(48.13) |
| No |  |  |  | 208(51.87) |
| Level of education (n=401) |  |  |  |  |
| Under certificate level |  |  |  | 173(43.14) |
| Certificate level |  |  |  | 120(29.93) |
| Degree and above |  |  |  | 108(26.93) |
| Religion (n=401) |  |  |  |  |
| Christian |  |  |  | 106(26.433) |
| Moslem |  |  |  | 122(30.424) |
| Not sure |  |  |  | 173(43.14) |
| Abbreviations: ANC –Antenatal care, SD – Standard deviation, Min-minimum value, Max-maximum value, (%) -percentage, (obs=401) number of observation |  |  |  |  |

### The measures of the level of acceptability of the 8-ANC visits among pregnant women

Findings from a One-sample t-test analysis indicate that the mean ANC contacts pregnant women could complete were 4-ANC contacts at SD of 1.9 contacts; at 95% CI [3.811, 4.184], P-value = 0.001, the findings significantly less than the 8-ANC contacts that World Health Organization recommends(29). The results further indicate that the overall utilization and acceptability rate among pregnant women to complete 8 or more ANC contacts was 24(5.99%), this proportion is unacceptably lower than expected. The high level of acceptability was mostly seen in the primigravida group at 27(6.73%) compared to those in the multigravida group at a low rate of 19(4.74%), with a regression coefficient of (-.222, at 95% CI [-.328 -.116], P=0.001), which suggests that has the number of pregnancies a woman has carried increases, it decreases the number of ANC contacts a pregnant woman could accept to attend by 22.1%. The disagreement to complete all the 8-ANC contacts was most seen in the multigravida group at a rate of 296(73.82%) which was significantly higher than in the primigravida group at 212(52.87**%)**. (Table 2) has the details

**Table 2.** Measures of the level of acceptability of the 8-ANC visits in pregnant women in their various categories.

**Indicators of acceptability of the 8-ANC visits**
| <u>Variables</u> | <u>ANC visits completed</u> |  | <u>Pregnant women completed the 8-ANC visits</u> |  |
| --- | --- | --- | --- | --- |
| 8- ANC contacts in weeks (n=401) | Mean (SD) of 8-ANC visits completed | 95% CI | The proportion of pregnant women who attended ≥ 8-ANC (%) | The pooled overall level of acceptability of 8-ANC visits (%) |
| Contact 1. |  |  |  |  |
| Contact 2 |  |  |  |  |
| Contact 3 |  |  |  |  |
| Contact 4 | 4(1.9)* | (3.80, 4.19)* |  |  |
| Contact 5 |  |  |  |  |
| Contact 6 |  |  |  |  |
| Contact 7 |  |  |  |  |
| ≥ Contact 8 | | | 24(5.99%) | 24(5.99%) |
| **Primigravida (n=401)** |  |  |  |  |
| Disagree to complete all 8-ANC |  |  | 212(52.87%)<sup>F</sup> |  |
| Somehow agree to complete 8-ANC |  |  | 162(40.40%)<sup>F</sup> |  |
| Strongly agree to complete 8-ANC |  |  | 27(6.73%)<sup>F</sup> |  |
| **Multigravida (n=401)** |  |  |  |  |
| Disagree to complete 8-ANC |  |  | 296(73.82%)<sup>F</sup> |  |
| Neutral |  |  | 86(21.45%)<sup>F</sup> |  |
| Strongly-agree to complete 8-ANC |  |  | 19(4.74%)<sup>F</sup> |  |
| * Mean (SD) is the mean and standard deviation), the CI, is Confidence interval and obtained from t test,<sup>F</sup>proportions and percentages obtained from Fisher's exact test. 8-ANC (%) is the 8-times Antenatal care visits, and percentage. |  |  |  |  |

**Table 3.** the best predictor variable to the acceptability and utilization of the 8-ANC visits from Pearson correlation coefficient (r) analysis.

| Predictor variables | Pearson correlation |  | P-value |
| --- | --- | --- | --- |
|  | coefficient(r) | R<sup>2</sup> |  |
| Age of pregnant women | 24.8(6.31)* | (24.29, . 27)* | <0.03 |
| Time of waiting in the queue (hours) | 5.34(5.09)* | (5.13, 5.58)* | <0.001 |
| Time ANC started (Gestation age in weeks) | 4.6(1.98)* | (4.35, 4.77)* | <.002 |
| Parity | 5.2(2.09) * | (4.02, 4.88) * | <0.03 |
| Distance to ANC clinic | 3.9(1.77)* | (3.70, 4.09)* | <0.001 |

ANC-antenatal care, \* Results from the Pearson correlation coefficient

**Table 4.** the predictors of the acceptability to complete the 8-ANC contacts among women attending ANC clinics in the Wakiso district.

| **Variables** | **Full Model 1** |  |  | **Nested model 2** |  |  |
| --- | --- | --- | --- | --- | --- | --- |
|  | **Estimates** | **P Value** | **95% Confidence interval** | **Estimates** | **P Value** | **95% Confidence interval** |
| Age(Years) | .003957 |  |  |  |  |  |
| Distance (KM) | .015325 |  |  |  |  |  |
| Time of waiting in queue(hours) | -.013468 |  |  | -.03843 | <0.0031** | (-.0728, -.00343) ** |
| Time ANC started (Gestation age in months) |  |  |  | -.15298 | <0.002** | (-.252 -.0538) ** |
| Parity | -.00779 | 0.700 |  |  |  |  |
| Being a primigravida |  |  |  |  |  |  |
| Disagree to complete 8-ANC | ref | ref | ref |  |  |  |
| Somehow agree to complete | .00405 | 0.938 | (-.098 .106) | ***** | ***** | ***** |
| Strongly agree to complete | -.1686 | 0.124 | (-.384 .047) |  |  |  |
| Being a Multigravida |  |  |  |  |  |  |
| Strongly disagree to complete 8-ANC | ref | ref | ref |  | ***** | ***** |
| Neutral on 8-ANC | -1.0278 | 0.000 | (-1.471 -.5839) | ***** |  |  |
| Strongly agree to complete | -.89172 | 0.041 | (-1.748 -.0346) |  |  |  |
| Level of income at home |  |  |  | 2.0246 | <0.000 ** | (3.001 4.047) ** |
| Low influence to complete 8-ANC | ref | ref | ref |  |  |  |
| Moderately influence to complete 8-ANC | -1.7318 | 0.130 | (-3.973 .5101) |  |  |  |
| Highly influence to complete | -2.7718 | 0.072 | (-5.788 .2445) |  |  |  |
| Level of education |  |  |  |  |  |  |
| Under-Certificate –level | ref | ref | ref |  |  |  |
| Certificate level | .06556 | 0.216 | (-.0383 .1695) | ***** | ***** | ***** |
| Degree and above | -.18244 | 0.143 | (-.4266 .0617) |  |  |  |
| Awareness about 8-ANC |  |  |  |  |  |  |
| No satisfactory information | ref | ref | ref | 1.4134 | <0.000** | (1.998 2.828) ** |
| Satisfactory information | .11509 | 0.043 | (.00374 .2264) |  |  |  |
| Very satisfactory information | -.25064 | 0.000 | (-.3794 -.1218) |  |  |  |
| Midwife motivation |  |  |  |  |  |  |
| Strongly demotivate | ref | ref | ref | ***** | ***** | ***** |
| Moderately demotivate | .10485 | 0.622 | (-.312 .522) |  |  |  |
| Highly motivate to complete | .19122 | 0.540 | (-.419 .802) |  |  |  |
| Attitude of Health work |  |  |  |  |  |  |
| Poor attitude of health workers |  |  |  |  |  |  |
| Somehow good attitude | ref | ref | ref |  |  |  |
| Very good attitude | .35916 | 0.429 | (-.5314 1.2498) | ***** | ***** | ***** |
|  | 1.6019 | 0.059 | (-.0584 3.2623) |  |  |  |

\*\* indicates significance at the 95% level \*\*\*\*\* Negligible Non-significant values, 95% level, GLM-generalized leaner model

### The inner setting and individual-level predictors on the acceptability and utilization of the 8-ANC contacts

The correlations and covariance analyses indicate that the level of acceptability to complete the 8-ANC contacts or more among pregnant women was significantly associated with the waiting time in the queue to be attended to by a midwife, *r* = -.70, *P <*0.001, and this was a moderate relationship. These findings suggest that a unit increase in hours of waiting in the queue to be served by a midwife decreases the number of ANC contacts a pregnant woman could attend at the ANC clinic. Results further indicate that Gestational Age (in weeks) at ANC contacts started, and the age of the pregnant woman were strongly correlated with the level of acceptability to complete the 8-ANC contacts, and these relationships were statistically significant, *r = -*.80, *p<*0.002, *r = -*.84, *p<*0.03, respectively. These findings suggest a unit increase in the age of a pregnant woman by years, and an increase in Gestational age by week, decreases the number of ANC contacts significantly

### The predictors of acceptability to complete the 8-ANC visits among pregnant women

Generalized leaner model (GLM)poison was fitted and indicated that the average waiting time in the queue to be served by a midwife was -.0384%, 95% CI; (-.073, -0343), and it was statistically significant (p-value= 0.003). The results suggest that a unit decrease in waiting time by an hour; increases the level of acceptability by 38.4% to complete the 8-ANC contacts or more. Further, a one-week gestation age decrease at which ANC started increases the acceptability probability to complete 8-ANC contacts by .153%, 95% CI; (-.252 -.054), and this increase was statistically significant (*P-*value *=*0.002). This signifies that one week decrease in gestation age at which a woman starts ANC contacts increases the level of acceptability to complete the 8-ANC contacts among pregnant women. The level of income influenced the acceptability of the 8-ANC contacts by 2.025%, 95% CI; (3.001 1.047), and it was statistically significant (*P-*value *=*0.001). The results suggest a unit increase in the level of income and an increase in the level of acceptability of the 8-ANC contacts among women attending to ANC clinic. The level of awareness about the completion of the 8-ANC contacts was also a potential predictor of acceptability women attended ANC clinic at 1.413%, 95%, CI; (1.998 3.828), and it was statistically significant (***P****-*value*=*0.001), and it suggests that as the level of awareness about the completion of 8-ANC contacts increases, increases the level of acceptability to complete all the 8-ANC contacts or more by 1.413%.

## Discussion

Maternal, perinatal deaths, stillbirth, and neonatal mortality rates are unacceptably high in low and middle-income countries (LMICs) Uganda inclusive. These deaths are a result of severe pregnancy complications which are preventable. It is a solemn public health concern, and to mitigate its escalation, WHO increased the number of antenatal visits from four (4) contacts to Eight (8) antenatal care visits “the 8-ANC contacts. However, there is a concern that pregnant women in Uganda do not complete all Eight (8) antenatal care contacts or more(7, 8, 16). Previous studies demonstrated that pregnant women don’t complete the 8-ANC visits, and the majority are noncompliant and nonadherence to the 8-ANC visits model(31-33)^5,26^. Unfortunately, scanty studies have assessed the implementation outcome on the acceptability of the 8-ANC visits model in Uganda. We performed a study design to test the hypothesis that the level of acceptability of the 8-ANC visits model among pregnant women in the Wakiso district is high and is associated with the completion of 8 or more ANC visits.

We found out that the pooled overall level of acceptability to complete the Eight (8) antenatal care contacts was significantly low as 24(5.99%) out of the 401 women participated in the study. The finding was relatively lower compared to the WHO antenatal care randomized trial for the evaluation of the new 8-ANC contact model of routine antenatal care established; and from other studies done with similar objectives(34-36). The results indicate a consistent trend across the disagreement interval scale to accept to complete all the 8-ANC contacts among pregnant women which resulted in a low level of acceptability of the 8-ANC contacts. It was established that a low level of acceptability was associated with the incompletion of the 8-ANC contacts among women attending ANC clinics in the Wakiso district. This indicates that the majority of pregnant women have not attended the ANC 8 times or more, as recommended by WHO(1), and as it was established by Odusina and coworkers(4, 37). ‘The high level of the unacceptability of Eight (8) antenatal care contacts or more was relatively higher among multigravidas than in primigravidas. There is an evidence gap from the previous studies to compare our research findings, and that makes our research findings novel. Furthermore, it was established that studies that have studied utilization, adoption, and other issues surrounding the completion of the 8-ANC contacts are scanty in Uganda’s context, and those with close contextual factors have got population gaps, different population characteristics, study settings, and different methodological approach (38-40)hence makes the study results different. Furthermore, other previous studies have assessed the acceptability of the 8-ANC model in high-resource setting countries, which had different populations of different health-seeking behaviour, and healthcare service delivery is at its prime. While other studies have assessed the acceptability of the 8-ANC model using insufficient sample sizes; consequently, the discrepancies are seen in the results of the WHO antenatal care randomized trial(34). This contributed to the disparities seen in many previous studies reviewed(4, 37).

The study further established the predictors of the acceptability to complete the Eight (8) Antenatal care contacts among pregnant women in Wakiso district, and we found out that, long waiting time in the queue to be attended to by a midwife, delays to start ANC “(gestation age at which ANC started)”, the level of income a pregnant woman has lack of awareness about the need to complete the 8-ANC contacts were the most statistically significant predictor variables influencing the acceptability to complete the 8-ANCvisits.; [.0384%, 95% CI; (-.073, -0343)], [.153%, 95% CI; (-.252 -.054)], [. 2.025%, 95% CI; (3.001 1.047)], [1.413, 95% CI; (1.998 3.828)], respectively.

The paradigm of our findings is that the 8-ANC contact model was not accepted in the social community, because of the established predictors in this study. Furthermore, the thinking is that this unacceptability was due to unforeseen internal and external contextual factors surrounding the implementation processes of the entire 8-ANC program in the community. For example, studies investigating the implementation outcome ‘**Feasibility** of the 8-ANC model, and evaluating the **appropriateness** of the 8-ANC model (41), using Rogers’ theory of innovation(42) are scanty. Otherwise, a feasibility study was not done to guide the implementation process of the new intervention. Diffusion of innovation guarantees program implementers, and program providers to evaluate the new intervention across the innovation characteristics(42) before it is scaled up in the community. This was not done, no wonder why acceptability of the 8-ANC visit model was low. Therefore, further study is needed to evaluate the feasibility and appropriateness of the 8-ANC contact model in the context of a low and mid-income country setting using a diffusion of innovation theory approach(42).

Our study has several strengths; The study had a large population turn-out, excellent control and sampling of participants, and multiple adjustments in the analysis. However, the study also had several limitations which include the use of observational study design which is the founder of confounders(43)

Taken together, our findings demonstrated that the overall level of acceptability to complete the Eight (8) antenatal care contact model is low among pregnant women in the Wakiso district, and this decline was mostly observed in multigravidas compared to the primigravidas. Furthermore, we have established that the long waiting time in the queue to be attended to by a midwife, the delay to start an ANC visit (high gestation age at which ANC started), the lack of community awareness about the 8-ANCvisits, and the level of income a pregnant woman attains were the predictor variables associated with the acceptability to complete the 8-ANC visits. Further, this study proves the existence of both internal and external contextual implementation challenges associated with the 8-ANC implementation processes that originated at the program design level to the implementation level. Additionally, the study guides the implementation approaches and steps that should be taken to scale up a new intervention in the social community. Hence the need for further study to evaluate the implementation outcome “feasibility” and “appropriateness” of the 8-ANC contact model in the context of a low and middle-income country setting using a diffusion of innovation theory approach is crucial.

## Data Availability

All data produced in the present work are contained in the manuscript

## Acknowledgment

The authors appreciate and supported the 8-ANC visit model acceptability project, for approval and access to the original data set.

